# Physical activity, mental health and well-being of adults during early COVID-19 containment strategies: A multi-country cross-sectional analysis

**DOI:** 10.1101/2020.07.15.20153791

**Authors:** James Faulkner, Wendy J. O’Brien, Bronagh McGrane, Daniel Wadsworth, John Batten, Christopher D. Askew, Claire Badenhorst, Erin Byrd, Maura Coulter, Nick Draper, Catherine Elliot, Simon Fryer, Michael J. Hamlin, John Jakeman, Kelly A. Mackintosh, Melitta A. McNarry, Andrew Mitchelmore, John Murphy, Helen Ryan-Stewart, Zoe Saynor, Mia Schaumberg, Keeron Stone, Lee Stoner, Beth Stuart, Danielle Lambrick

## Abstract

**Objectives:** To assess how the early stages of National governments Coronavirus disease (COVID-19) containment strategies impacted upon the physical activity, mental health and well-being of adults in the UK, Ireland, New Zealand and Australia

**Design:** Observational, cross-sectional

**Setting:** Online survey disseminated in the UK, Ireland, New Zealand and Australia within the first 2-6 weeks of government mandated COVID-19 restrictions

**Participants:** Adults (n = 8,425; 44.5 ± 14.8 y), ≥ 18 y who were residing in the surveyed countries

**Main outcome measures:** Stages of Change scale for exercise behaviour change, International Physical Activity Questionnaire (short-form), World Health Organisation-5 Well-being Index and the Depression Anxiety and Stress Scale-9

**Results:** Participants who reported a negative change in exercise behaviour between pre- and during the early COVID-19 restrictions demonstrated poorer mental health and well-being compared to those who had either a positive change- or no change in their exercise behaviour (*p*<0.001). Whilst women reported more positive changes in exercise behaviour, young people (18-29y) reported more negative changes (both *p*<0.001). Individuals who engaged in more physical activity reported better mental health and well-being (*p*<0.001). Although there were no differences in physical activity between countries, individuals in New Zealand reported better mental health and well-being (*p*<0.001).

**Conclusion:** The COVID-19 restrictions have differentially impacted upon the physical activity habits of individuals based upon their age and sex, and therefore have important implications for international policy and guideline recommendations. Public health interventions that encourage physical activity should target specific groups (e.g., men, young adults) who are most vulnerable to the negative effects of physical distancing and/or self-isolation.

## Introduction

At the onset of the novel coronavirus disease (COVID-19) pandemic, governments in various countries implemented national containment strategies to limit the spread of the virus and reduce the risk of national healthcare systems becoming critically overburdened. Early containment strategies have included: government-mandated self-isolation for those in ‘high-risk’ groups (e.g., ≥ 70 y), and a 14-day isolation period for individuals showing symptoms or receiving a diagnosis of COVID-19; closed borders and travel restrictions; closures of schools and childcare providers; 2m physical distancing measures; and closures of services and amenities (gyms, pools, etc.). Although physical distancing and self-isolation regulations aim to reduce person-to-person transmission of COVID-19, there are potentially significant public health implications from such measures, such as poor lifestyle behaviours (e.g., reduced physical activity [PA]), impaired physical and psychological health, and higher all-cause mortality.[1]

Physical activity is known for its beneficial effects on the immune system and for counteracting many comorbidities, such as obesity, diabetes, hypertension, and mental health disorders.[1, 2] Under non-pandemic circumstances, modern lifestyle behaviours (e.g., excessive screen time) encourage physical *inactivity* and sedentariness,[3] but whether this is exacerbated by containment strategies during COVID-19 is unknown. Many opportunities to be physically active, such as participation in community- or hospital-based rehabilitation programmes, and use of fitness centres and public parks have been prohibited or restricted for people of all ages as a result of the physical distancing and self-isolation directives. Incidental PA has likely reduced for a substantial proportion of the population worldwide due to the increased number of people ‘working from home’ or ‘out of work’ as a direct result of COVID-19. Indeed, early reports from the United States of America (USA) suggest that among individuals who met PA guidelines prior to COVID-19, those engaging in physical distancing showed a 32% decrease in PA during early COVID-19 restrictions.[4] Furthermore, individuals who did not meet recommended PA guidelines and engaged in greater screen time presented with higher depressive symptoms and stress than those who were more physically active.[4]

Although containment strategies may have introduced new barriers to being physically active for some, a change in work and social patterns may have facilitated additional opportunities to engage in PA for others. For example, an increase in available time (e.g., reduced commute time) and access to various free online exercise classes (e.g., yoga/Pilates, high intensity interval training [HIIT]), may have provided individuals with opportunities to maintain or increase their PA during early COVID-19 restrictions. Indeed, despite strict government regulations in some countries, ‘daily exercise’ was one of the few reasons people could leave their homes and this may have been an incentive for some people to increase their PA.

While the types of COVID-19 restrictions implemented have been broadly similar globally, the timing and enforcement of these have differed considerably across countries. Differing government approaches likely contributed to the significant differences in COVID-19 infection and death rates (e.g. between UK, Ireland [IRE], New Zealand [NZ], and Australia [AUS]), and may also contribute to differences in the behavioural, physical and mental health impacts of the pandemic which are yet to be explored. Accordingly, the purpose of this study was to assess PA, mental health and well-being during early COVID-19 restrictions across the UK, IRE, NZ and AUS populations. It was hypothesised that individuals who were physically active during COVID-19 restrictions would demonstrate better mental health and well-being than those who were not.

## Methods

### Study design

This study was designed to collect cross-sectional data using online survey’s during the early government-led COVID-19 containment strategies (April and May, 2020). The overall programme of research, which also includes longitudinal components, was approved by the Faculty of Health and Wellbeing Ethics Committee, University of Winchester, UK (HWB/REC/20/04). This study adhered to Strengthening the Reporting of Observational Studies in Epidemiology (STROBE) guidelines.[5]

### Population sampling

Sampling commenced between 10 days and 6 weeks of government mandated COVID-19 restrictions (Table 1). Convenience sampling using mass emailing via collaborating author networks, social media (Twitter, Facebook) and mass media engagement (radio, newspapers), and snowball sampling, were used to recruit participants. Adults (≥18 y) who were residing in the surveyed countries were eligible to participate. All participants provided informed consent at the start of the survey.

**Table 1:**
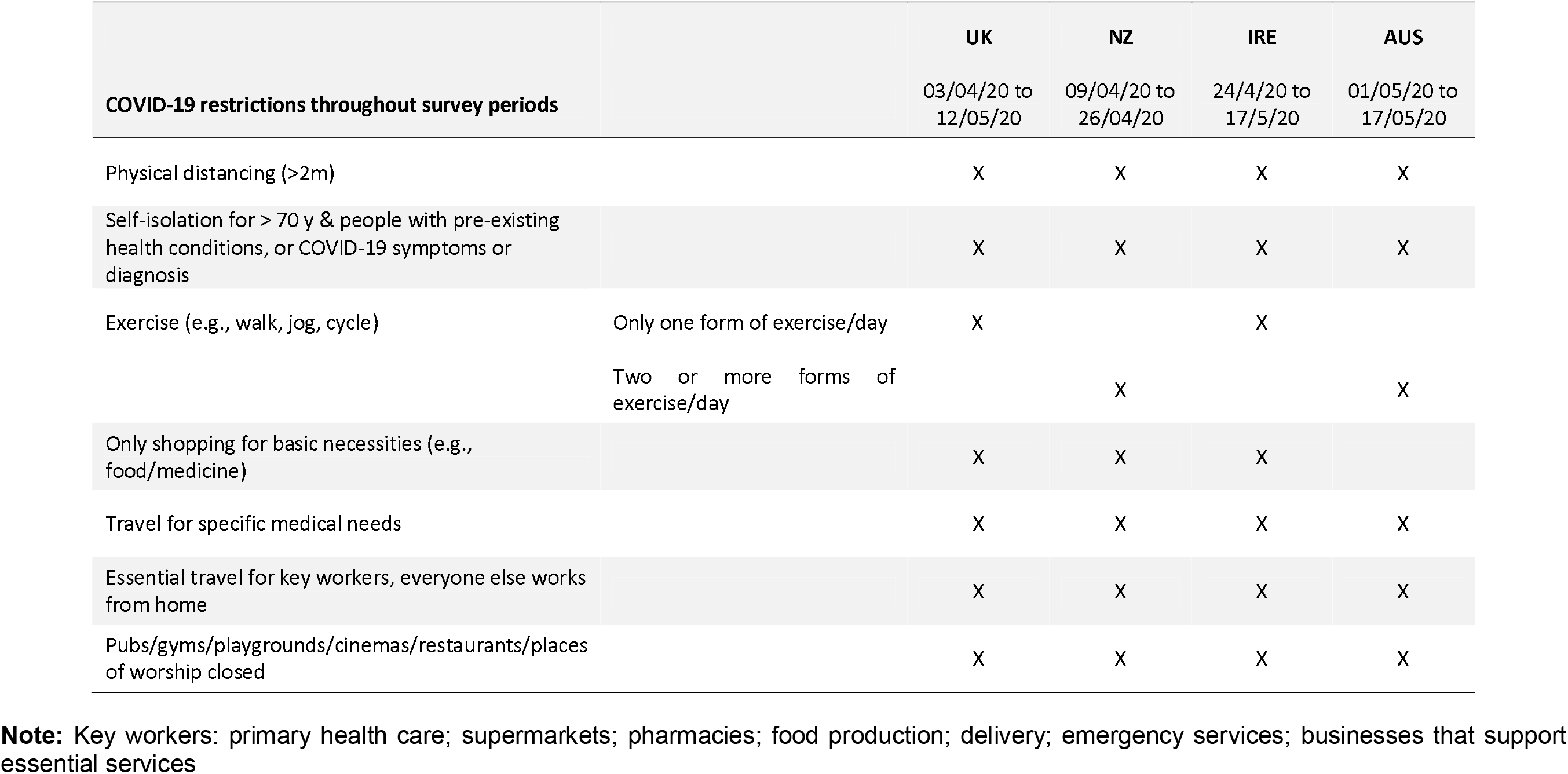
COVID-19 restrictions to human movement

### Survey and data collection

The survey was administered using JISC (Bristol, UK) or Qualtrics (London, UK) and was designed to take no longer than 15 minutes to complete. Participants self-reported demographic information and completed questionnaires relating to PA (the International Physical Activity Questionnaire: Short Form [IPAQ-SF]),[6] exercise behaviour change (Stages of Change scale),[7] mental health (Depression Anxiety and Stress Scale-9 [DASS-9]),[8] and well-being (World Health Organisation-5 Well-being Index [WHO-5]),[9] and described their weekly PA using free-text responses. All measures were assessed during the early COVID-19 restrictions with the exception of the Stages of Change scale and weekly PA free-text responses, which also captured pre-COVID-19 restriction information. In addition, participants reported whether they met recommended guidelines for daily PA (≥150 minutes of moderate-to vigorous intensity PA each week) before the COVID-19 restrictions were imposed.[10]

### Survey Measures

#### Physical activity

A Stages of Change scale was used to capture participant intentions and involvement in the exercise behaviour change process.[7] Participants self-reported their pre- and during COVID-19 exercise behaviour based on one of the following statements: i) I currently do not exercise and do not intend to start in the next 6 months; ii) I currently do not exercise but I am thinking about starting in the next 6 months; iii) I currently exercise a little but not regularly; iv) I currently exercise regularly but have begun doing so in the last 6 months; or v) I currently exercise regularly and have done so for more than 6 months. These statements correspond with the Pre-contemplation, Contemplation, Preparation, Action, and Maintenance Stages of Change of the Transtheoretical Model of Behaviour Change, respectively. In addition, weekly PA (i.e., type) pre- and during early COVID-19 restrictions were recorded as free-text questions in the survey, with participants also completing the IPAQ-SF.

The IPAQ-SF consists of seven questions asking individuals to recall the previous week’s PA (days per week, total minutes per day), with regards to walking, and moderate- and vigorous-intensity activities, and average daily sitting time. The IPAQ-SF is an acceptable tool to assess PA in large population samples across various age groups (e.g., 18-70 y),[6] and has been validated (*r* = 0.67) and tested for reliability (*rho* = 0.77-1.00) across many countries.[11]

#### Mental health and well-being

The DASS is a widely used self-reported scale that concurrently assesses symptoms of depression, anxiety, and stress. This study used the DASS-9, an empirically derived version based on the DASS-21.[8] This nine-item questionnaire consists of three subscales (depression, anxiety and stress) with three items each. The WHO-5 was used as a short global rating scale to measure subjective well-being. It has been demonstrated to have good contrast validity and can be used in both clinical practice and in research studies.[9] The WHO-5 includes the following items: i) ‘I have felt cheerful and in good spirits’, ii) ‘I have felt calm and relaxed’, iii) ‘I have felt active and vigorous’, iv) ‘I woke up feeling fresh and rested’ and v) ‘My daily life has been filled with things that interest me’.

### Data analysis

For the Stages of Change scale, changes in exercise intentions and behaviours results were reported as no change, positive change (increased rating from pre-to during COVID-19 restrictions), or negative change (decreased rating from pre-to during early COVID-19 restrictions).

For the IPAQ-SF, results were reported as a continuous variable (MET·min^−1^·week^−1^) and in categories (low-, moderate- or high-PA levels). For each activity (walking, moderate, vigorous), the amount (days per week) and duration (minutes per day) of PA was multiplied by an appropriate MET level (1 MET = 3.5 mL·kg^−1^·min^−1^) to determine MET·min^−1^·week^−1^.[7, 12] IPAQ-SF categories included: high (seven or more days of any combination of walking, moderate- or vigorous-intensity PA achieving ≥3,000 MET·min^−1^·week^−1^), moderate (five days or more of any combination of walking, moderate- or vigorous intensity PA achieving ≥600 to 2999 MET·min^−1^·week^−1^) and low (achieving □600 MET·min^−1^·week^−1^) levels of PA.

Each of the five items of the WHO-5 were scored from 0 to 5. The total raw score was translated into a percentage scale ranging from 0 (absence of well-being) to 100 (maximal well-being). All nine items of the DASS-9 were scored on a scale from 0 (none of the time) to 3 (most of the time). As such, the three items of the DASS-9 (depression, anxiety and stress) were each cumulatively scored between 0 and 9, with higher scores demonstrating poorer mental health.

Free-text PA was thematically coded by collaborating authors based upon the Compendium of Physical Activity,[12] accounting for the type of activity in which participants engaged. Data was originally coded into 32 activity categories, and then aggregated into 13 higher level activity groupings (Supplementary Table 1). “Online” activity was categorised and included non-face-to-face activities (YouTube videos, Zoom, etc.). Coding was checked by JF and W’OB.

### Statistical analysis

The analysis was primarily descriptive, with proportions reported for binary and categorical variables and means and standard deviations or medians and interquartile ranges reported as appropriate to the distribution of the continuous variables. For the IPAQ classification, between group differences were explored using chi-squared tests. To explore changes in physical activity levels, multinomial logit models were used. For WHO-5, DASS-9 and IPAQ-SF, multivariable linear regression was used to obtain the independent effect of each characteristic on the outcome. Spearman’s correlation coefficient (*rho*) was used to quantify the association between PA with mental health and wellbeing measures. Statistical analysis was completed on Stata (version 16).

## Results

Of the 8,425 participants recruited (Figure 1), 3,121 were residing in the UK, 4,007 in NZ, 903 in IRE and 394 in AUS. Only individuals who completed all survey items were included in the statistical analysis. Respondents were predominantly white females (Table 2). Data for exercise behaviours, IPAQ-SF, WHO-5 and DASS-9, pre- and/or during early COVID-19 restrictions are presented in Table 3.

**Table 2:** Population characteristics by country and in total.

|  | UK | NZ | IRE | AUS | Total |
| --- | --- | --- | --- | --- | --- |
| **Sample (n)** | 3,121 | 4,007 | 903 | 394 | 8,425 |
| **Sex (n/%)** |  |  |  |  |  |
| Male | 1,024 (32.8%) | 1,087 (27.1%) | 232 (25.7%) | 89 (22.6%) | 2,432 (28.9%) |
| Female | 2,094 (67.1%) | 2,886 (72.0%) | 668 (74.0%) | 304 (77.2%) | 5,952 (70.7%) |
| Prefer not to say | 3 (0.1%) | 24 (0.9%) | 3 (0.3%) | 1 (0.3%) | 31(0.5%) |
| **Mean age (y) (x/SD)** | 43.9 (14.9) | 46.5 (14.7) | 38.7 (13.3) | 41.1 (14.0) | 44.5 (14.8) |
| **Age Groups (n/%)** |  |  |  |  |  |
| 18-29 y | 627 (20.2%) | 619 (15.5%) | 252 (27.9%) | 103 (26.1%) | 1,601 (19.0%) |
| 30-39 y | 722 (23.2%) | 775 (19.3 %) | 248 (27.5%) | 103 (26.1 %) | 1,848 (22.0%) |
| 40-49 y | 650 (20.9%) | 910 (22.7%) | 205 (22.7%) | 86 (21.8%) | 1,851 (22.0%) |
| 50-59 y | 559 (18.0%) | 853 (21.3%) | 132 (14.6%) | 55 (14.0%) | 1,599 (19.0%) |
| 60-69 y | 392 (12.6%) | 578 (14.4%) | 50 (5.5%) | 36 (9.1%) | 1,056 (12.6%) |
| 70-79 y | 141 (4.5%) | 250 (6.2%) | 16 (1.8%) | 8 (2.0%) | 415 (4.9%) |
| 80+ y | 15 (0.5%) | 22 (0.5%) | 0 (0.0%) | 3 (0.8%) | 40 (0.5%) |
| **Ethnicity (n/%)** |  |  |  |  |  |
| White | 3,010 (96.4%) | 3,630 (90.6%) | 894 (99.0%) | 361 (93.7%) | 7,895 (93.8%) |
| Polynesian | 0 (0.0%) | 156 (3.9%) | 0 (0.0%) | 0 (0.0%) | 156 (1.9%) |
| Indian | 37 (1.2%) | 48 (1.2%) | 4 (0.4%) | 8 (2.0%) | 97 (1.2%) |
| Asian | 0 (0.0%) | 95 (2.4%) | 0 (0.0%) | 2 (0.5%) | 97 (1.1%) |
| Black | 14 (0.5%) | 8 (0.2%) | 2 (0.2%) | 2 (0.5%) | 26 (0.3%) |
| Mixed groups | 37 (1.2%) | 18 (0.5%) | 2 (0.2%) | 1 (0.3%) | 58 (0.7%) |
| Other | 23 (0.7%) | 52 (1.3%) | 1 (0.1%) | 12 (3.1%) | 88 (0.9%) |
| **Household (pre- COVIS-19) (n/%)** |  |  |  |  |  |
| Live alone | - | 515 (12.9%) | 84 (9.3%) | 45 (11.4%) | 644 (12.1%) |
| Couple | - | 1,379 (34.4%) | 184 (20.4%) | 146 (37.1%) | 1,709 (32.2%) |
| Two parent family | - | 1,162 (29.0%) | 355 (39.3%) | 112 (28.4%) | 1,629 (30.7%) |
| Single parent family | - | 121 (3.0%) | 22 (2.4%) | 14 (3.6%) | 157 (3.0%) |
| Extended family | - | 328 (8.2%) | 146 (16.2%) | 33 (8.4%) | 507 (9.6%) |
| Shared occupancy | - | 501 (12.5%) | 112 (12.4%) | 32 (8.1%) | 645 (12.2%) |
| Residential care | - | 1 (0.02%) | 0 (0.0%) | 0 (0.0%) | 1 (0.02%) |
| **Household (during COVID-19) (n/%)** |  |  |  |  |  |
| Live alone | - | 459 (11.5%) | - | 72 (8.0%) | 531 (10.8%) |
| Couple | - | 1,287 (32.1%) | - | 184 (20.4%) | 1,471 (30.0%) |
| Two parent family | - | 164 (29.1%) | - | 366 (40.5%) | 1,530 (31.2%) |
| Single parent family | - | 114 (2.9%) | - | 28 (3.1%) | 142 (2.9%) |
| Extended family | - | 588 (14.7%) | - | 206 (22.8%) | 794 (16.2%) |
| Shared occupancy | - | 395 (9.9%) | - | 46 (5.1%) | 441 (9.0%) |
| Residential care | - | 0 (0.0%) | - | 1 (0.1%) | 1 (0.02%) |
| **Comorbidity (n/%)** | - | 873 (21.8%) | 173 (19.2%) | 15 (3.8%) | 1061 (20.0%) |

**Note:** Black includes African, Caribbean, African-American; Indian includes East Indian, Pakistani, Bangladeshi; Asian includes Chinese, Japanese and all South-East Asian; Polynesian includes Māori, Pacific Islanders

**Table 3:**
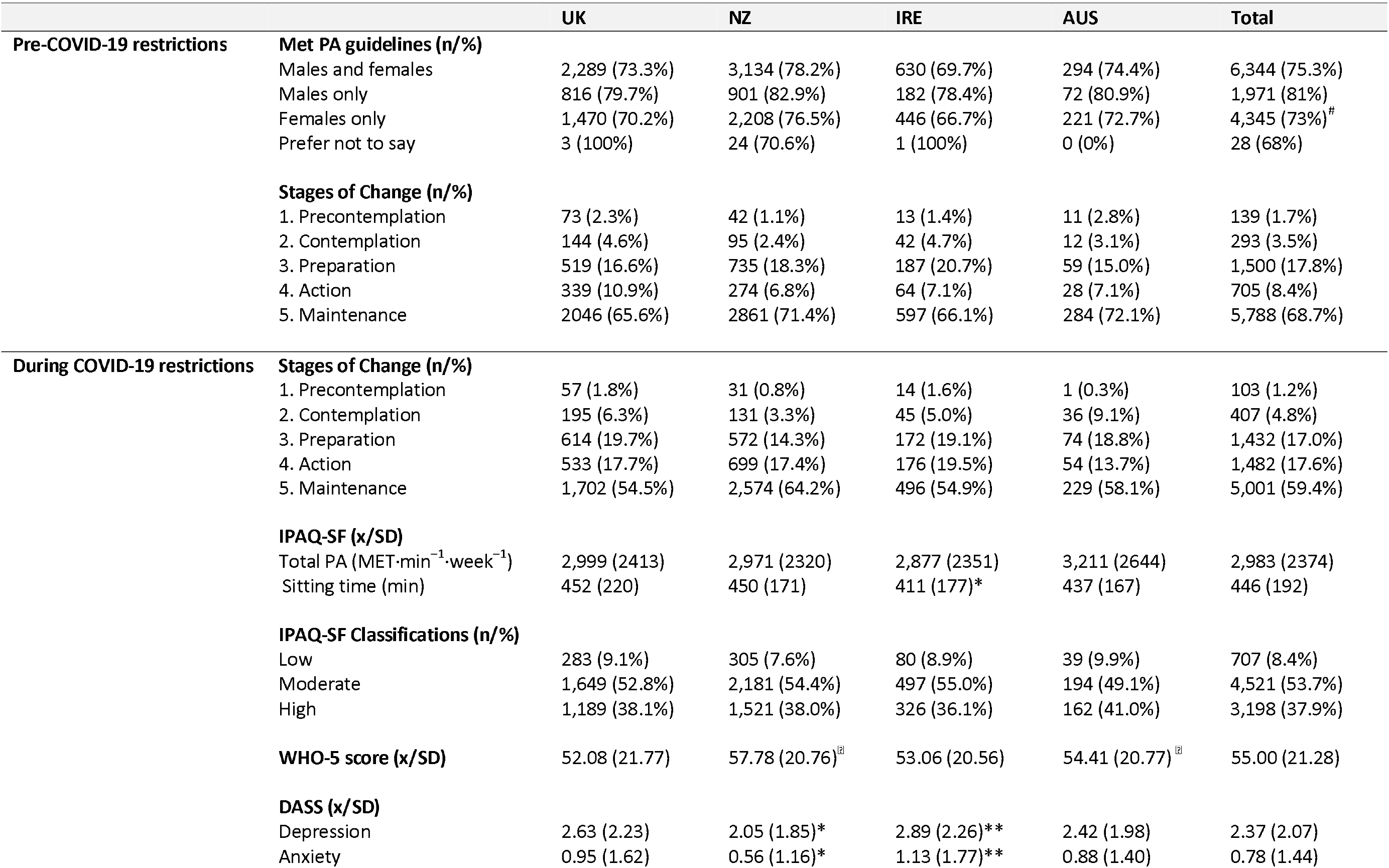
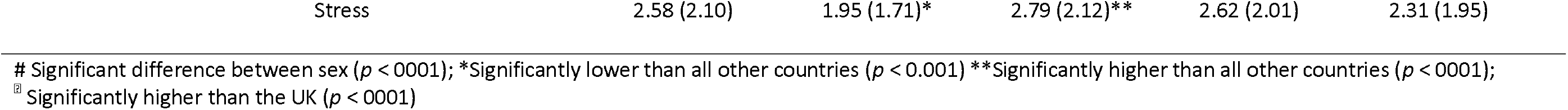
Pre- and during COVID-19 restrictions for physical activity instruments, WHO-5 and DASS

**Figure 1:**
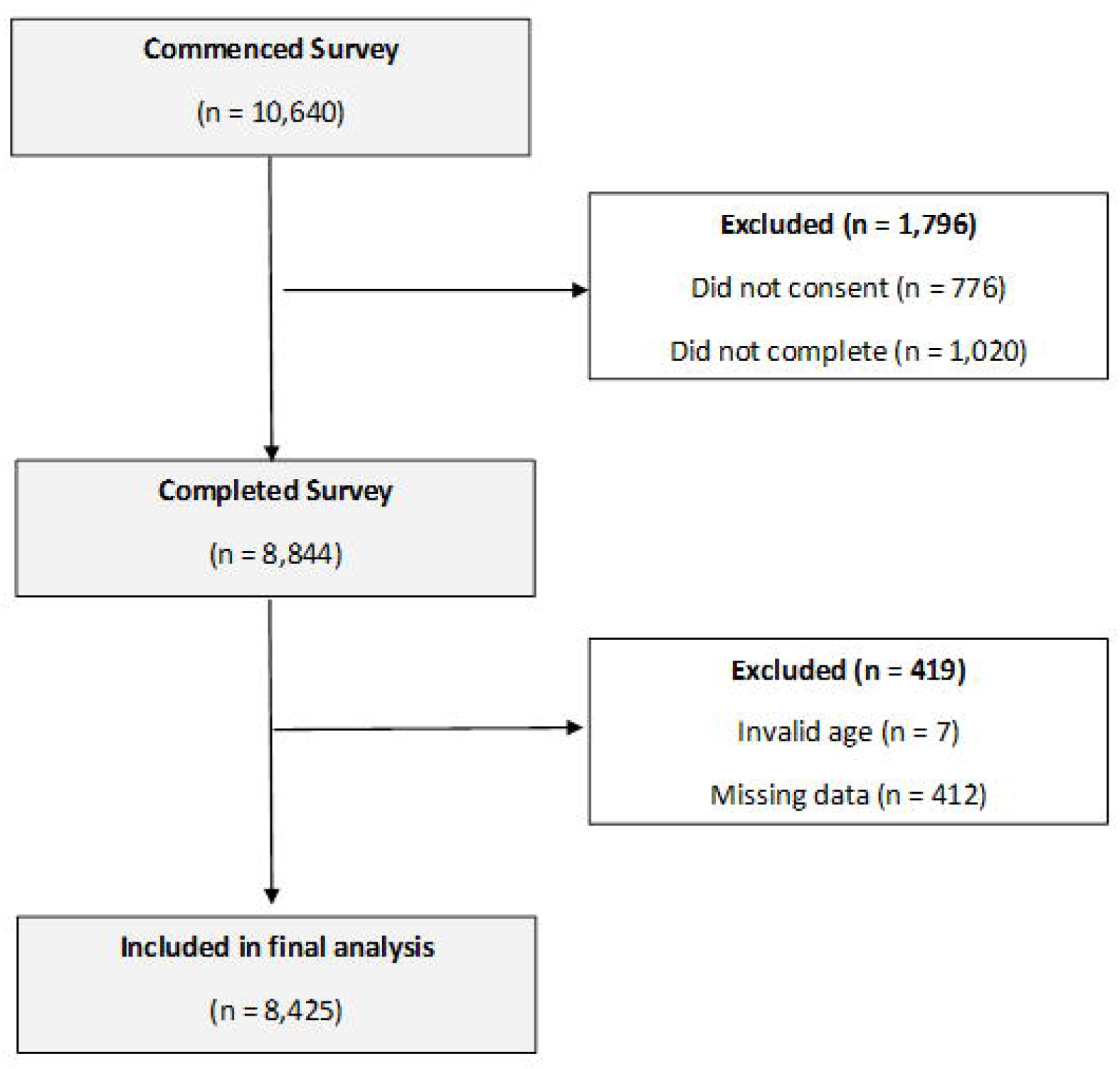
Flow diagram

### Physical activity, mental health and well-being

Fewer females met the recommended PA guidelines before COVID-19 restrictions compared to males (*p* < 0.001; 73% vs 81%, respectively; Table 3). During early COVID-19 restrictions, there were no differences in PA between countries (*p* > 0.05; Table 3), although females engaged in less high-intensity PA than males (*p* < 0.001; 36% vs 41%, respectively), irrespective of country. Sitting time was lower for IRE compared to all other countries (*p* < 0.001), with no differences between the UK, NZ and AUS (*p* > 0.05). Depression, anxiety and stress were lower in NZ compared to UK, AUS and IRE (*p* < 0.001), whereas IRE reported higher scores than all other countries (*p* < 0.001; Table 3). Well-being was higher in NZ and AUS than the UK (*p* < 0.001), but there was no significant difference between the UK and IRE (*p* > 0.05; Table 3).

Using combined data from all four countries, Spearman’s correlation coefficient (*rho*[95%CI]) demonstrated moderate positive correlations between PA and WHO-5 scores (*rho* = 0.35 [0.33, 0.37]) and negative correlations between PA and depression (*rho* = −0.24 [-0.26,-0.22]), anxiety (*rho* = −0.13 [-0.15,-0.11]) and stress (*rho* = −0.13 [-0.14,-0.10]). As sitting time increased, negative correlations were reported with the WHO-5 *(rho* = −0.20 [-0.22,-0.18]), while positive correlations were reported with depression (*rho* = 0.18 [0.16,0.20]), anxiety (*rho* = 0.08 [0.05,0.10]) and stress (*rho* = 0.08 [0.06,0.10]).

There was, on average, 32% change (positive or negative) in exercise behaviour pre-to during COVID-19 restrictions. There were differences between countries, with the UK and AUS reporting the greatest negative change in exercise behaviour (*p* < 0.001; Table 4). NZ demonstrated the least change in exercise behaviour as a result of the early COVID-19 restrictions. Further analysis demonstrated significant differences in exercise behaviour according to gender, age, and comorbidity (all *p* < 0.001; Table 5). Specifically, females reported more positive changes in their exercise behaviour compared to males, while younger people (18-29 y) reported more negative changes than other age groups during early COVID-19 restrictions. Individuals with self-reported comorbidities were more likely to make a change in their exercise behaviour than those without (*p* < 0.001), with a similar percentage reporting a positive or negative change in exercise behaviour. When adjusted for age, gender, and ethnicity, individuals who demonstrated a negative change in exercise behaviour had significantly higher DASS-9 scores and significantly lower WHO-5 scores compared to those who had either a positive change- or no change in their exercise behaviour (all *p* < 0.001; Table 6).

**Table 4:** Exercise behaviour change (positive, negative, no change) between pre- and during COVID-19 restrictions. Data presented as n (%)

| Exercise behaviour | UK | NZ | IRE | AUS | Total |
| --- | --- | --- | --- | --- | --- |
| Positive change | 466 (14.9%) | 609 (15.2%) | 153 (16.9%) | 52 (13.2%) | 1,280 (15.2%) |
| Negative change | 666 (21.3%)* | 505 (12.6%) | 160 (17.7%) | 89 (22.6%)* | 1,420 (16.8%) |
| No change | 1,989 (63.7%) | 2,893 (72.2%) | 590 (65.3%) | 253 (64.2%) | 5,725 (67.9%) |

\*Significantly greater negative change in exercise behaviour than NZ or IRE ( $p < 0001$ )

**Table 5:** Changes in exercise behaviours (positive, negative, no change) by sex, age groups and ethnicity.

|  | Negative change<br>(n=1420) | Relative risk compared<br>to no change | No change (n=5725) | Positive change<br>(n=1280) | Relative risk compared<br>to no change |
| --- | --- | --- | --- | --- | --- |
| **Sex** |  |  |  |  |  |
| Male | 397 (16.3%) | REF | 1,741 (71.6%) | 294 (12.1%) | REF |
| Female | 1,008 (16.9%) | 1.11 (0.98, 1.27) | 3,967 (66.7%) | 977 (16.4%)* | 1.46 (1.27, 1.68) |
| **Age group** |  |  |  |  |  |
| 18-29 | 418 (26.1%)* | REF | 946 (59.1%) | 237 (14.8%) | REF |
| 30-39 | 353 (19.1%) | 0.69 (0.58, 0.81) | 1,160 (62.8%) | 335 (18.1%) | 1.15 (0.96, 1.39) |
| 40-49 | 290 (15.7%) | 0.52 (0.44, 0.62) | 1,260 (68.1%) | 301 (16.3%) | 0.95 (0.79, 1.15) |
| 50-59 | 187 (11.7%) | 0.34 (0.29, 0.43) | 1,186 (74.2%) | 226 (14.1%) | 0.76 (0.62, 0.93) |
| 60-69 | 117 (11.1%) | 0.33 (0.26, 0.42) | 796 (75.4%) | 143 (13.5%) | 0.72 (0.57, 0.90) |
| 70-79 | 46 (11.1%) | 0.31 (0.22, 0.43) | 338 (81.5%) | 31 (7.5%) | 0.37 (0.25, 0.54) |
| 80+ | 5 (12.5%) | 0.35 (0.14, 0.91) | 32 (80.0%) | 3 (7.5%) | 0.37 (0.11, 1.23) |
| **Ethnicity** |  |  |  |  |  |
| White | 1,314 (16.6%) | REF | 5,406 (68.4%) | 1,183 (14.9%) | REF |
| Polynesian | 21 (13.5%) | 0.89 (0.55, 1.43) | 97 (62.2%) | 38 (24.4%)* | 1.79 (1.22, 2.61) |
| Indian | 26 (26.8%) | 2.02 (1.26, 3.24) | 53 (54.6%) | 18 (18.6%) | 1.55 (0.91, 2.67) |
| Asian | 15 (15.5%) | 0.91 (0.52, 1.59) | 68 (70.1%) | 14 (14.4%) | 0.94 (0.52, 1.68) |
| Black | 8 (30.8%) | 2.35 (0.98, 5.62) | 14 (53.9%) | 4 (15.4%) | 1.31 (0.43, 3.97) |
| Mixed | 17 (29.3%) | 2.33 (1.28, 4.24) | 30 (51.7%) | 11 (18.6%) | 1.68 (0.84, 3.35) |
| Other | 19 (21.6%) | 1.37 (0.81, 2.31) | 57 (64.8%) | 12 (13.6%) | 0.96 (0.51, 1.80) |
| **Comorbidity** |  |  |  |  |  |
| Yes | 183 (17.3%) | REF | 689 (64.9%) | 189 (17.8%) | REF |
| No | 456 (12.6%)* | 0.66 (0.54, 0.79) | 2,260 (72.6%) | 535 (14.8%)* | 0.74 (0.62, 0.90) |

\*Significantly difference between items ( $p < 0.001$ ). **Note:** REF, Reference value

**Table 6:** Mean (SD) WHO-5 and DASS-9 scores for positive, negative and no change in exercise behaviours. Mean difference (± 95% CI) reported when comparing no change and positive exercise behaviour with negative change in exercise behaviour

|  | Negative change | No change |  | Positive change |  |
| --- | --- | --- | --- | --- | --- |
|  | x (SD) | x (SD) | Mean difference compared to negative change (95% CI) | x (SD) | Mean difference compared negative change (95% CI) |
| **WHO-5 score** | 40.52 (19.97)* | 58.48 (20.45) | 16.2 (15.0, 17.3) | 55.53 (19.54) | 14.3 (12.9, 15.8) |
| **DASS-9** |  |  |  |  |  |
| Depression | 3.65 (2.39)** | 2.09 (1.89) | -1.3 (-1.5, -1.2) | 2.22 (1.94) | -1.3 (-1.5, -1.2) |
| Anxiety | 1.24 (1.85)** | 0.65 (1.30) | -0.5 (-0.5, -0.4) | 0.84 (1.44) | -0.3 (-0.4, -0.2) |
| Stress | 3.03 (2.21)** | 2.13 (1.85) | -0.7 (-0.8, -0.5) | 2.26 (1.92) | -0.7 (-0.8, -0.5) |

Significantly lower than either positive or no change in exercise behaviour ( $p < 0.001$ ) \*\*Significantly higher than positive or no change in exercise behaviour ( $p < 0.001$ )

The type of PA participants engaged in pre- and during early COVID-19 restrictions are presented in Figure 2 and Supplementary Tables 2 and 3.

**Figure 2:**
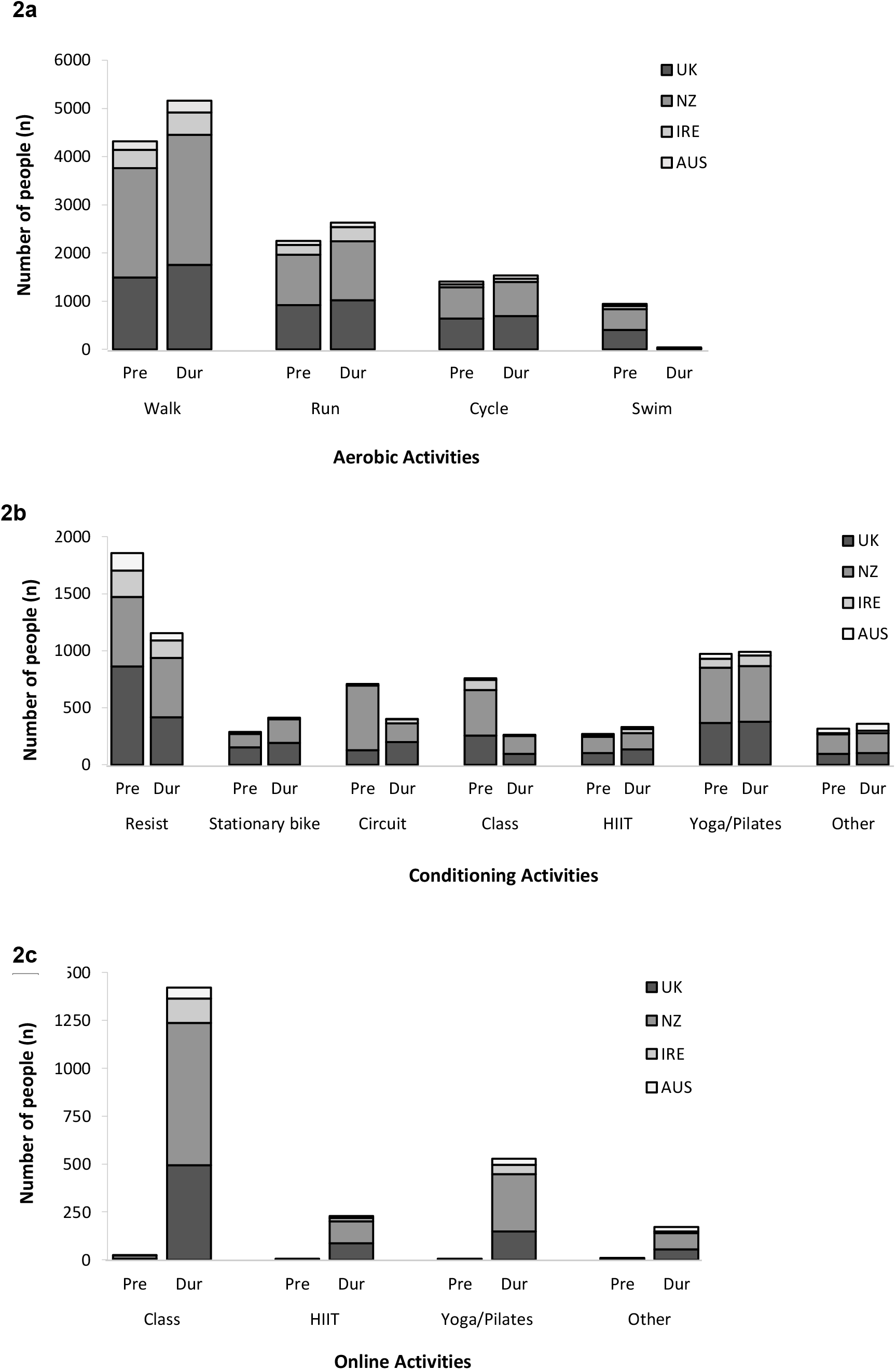
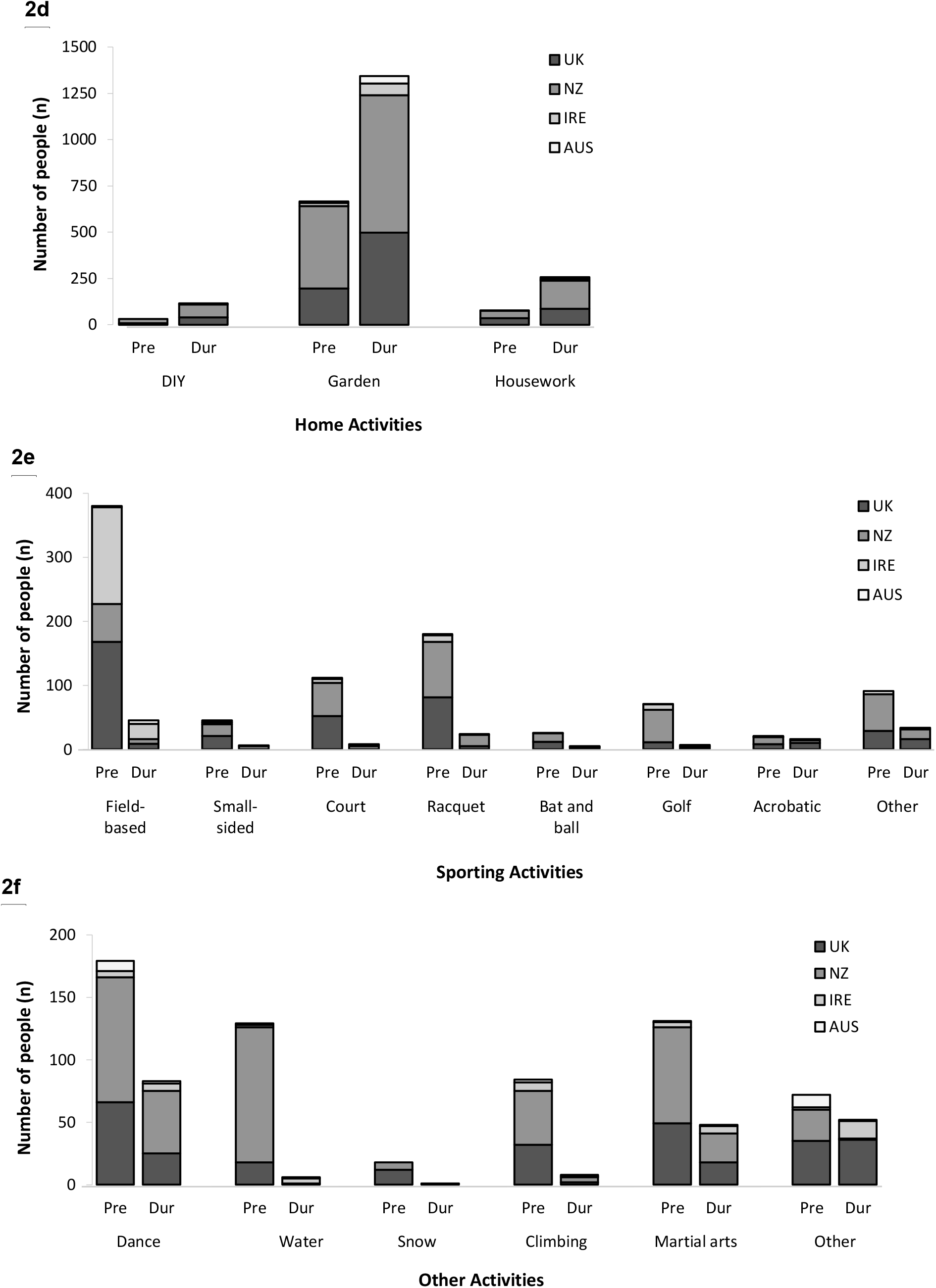
Number of people taking part in activities pre- and during COVID-19 restrictions: 2a) Aerobic activities, 2b) Conditioning activities, 2c) Online activities, 2d) Home activities, 2e) Sporting activities, and 2f) Other activities. ***Note:*** AUS, Australia; DIY, Do-It-Yourself; Dur, During COVID-19 restrictions; HIIT, High Intensity Interval Training; IRE, Ireland; NZ, New Zealand; Pre, Pre-Covid-19 restrictions; UK, United Kingdom

## Discussion

### Statement of principal findings

This study demonstrated that individuals who had a negative change in their exercise behaviour between pre- and during early COVID-19 restrictions reported poorer mental health and well-being; a relationship that was evident across all countries investigated. Females reported more positive changes in exercise behaviour compared to males, while younger people (18-29 y) reported more negative changes in exercise behaviour compared to all other age groups. There were no differences in the amount of PA people engaged in during COVID-19 restrictions across the four countries surveyed. There were differences, however, in mental health and well-being, with those in NZ reporting better outcomes than those in the UK, IRE or AUS. These findings have important implications for policy and guideline recommendations to encourage people to be physically active, and thus promote better mental health and well-being, throughout the COVID-19 pandemic and recovery period.

### Strengths and weaknesses of the study

The study findings should be contextualised in light of methodological limitations and strengths. The predominant ethnicity and sex of the respondents were white females which may not reflect the total population of the countries surveyed. Given that racial and/or ethnic disparities may impact the burden of COVID-19 related outcomes,[13] further investigation of the relationship between PA and mental health during COVID-19 in various racial and/or ethnic groups is needed. Furthermore, 75% of participants suggested that they met recommended PA guidelines of engaging in ≥150 minutes of moderate-to vigorous intensity PA each week; higher than the population average of countries surveyed.[14, 15] Although a wide and varied recruitment strategy was implemented, there may have been an inherent sampling bias due to the study authors working broadly within the fields of sport, exercise and health, thus the transferability to other population groups is less assured.

Strengths of this study include the sample size and the speed with which the surveys were implemented within all four countries. This ensured that the population response to the respective government-mandated containment strategies was captured at similar levels of restriction across all countries, and facilitated our planned longitudinal study design. A further strength is that all countries used identical primary outcome measures (IPAQ-SF, WHO-5, DASS-9, Stages of Change scale), thus enabling the pooling of data and between-country comparisons.

### Findings in relation to other studies

#### Exercise behaviour change

A potential implication of physical distancing is that poor lifestyle behaviours may be intensified.[2] However, the government containment strategies associated with the surveyed countries allowed individuals to engage in differing daily PA and/or exercise, which afforded the opportunity for people to meet the recommended guidelines of ≥150 minutes of moderate-to vigorous intensity PA each week. In this study, individuals (83% of sample) who reported no change or a positive change in exercise behaviour from pre-to during COVID-19 restrictions reported better mental health (lower DASS-9 scores) compared to individuals who reported a negative change in their exercise behaviour. Similarly, individuals who reported a negative exercise behaviour change exhibited a substantially lower WHO-5 score compared to people who reported no changes (95% CI: 15.0 to 17.3 points lower) or a positive change (95% CI: 12.9 to 15.8 points lower) in exercise behaviour. As the threshold for a clinically relevant change on the WHO-5 is 10 points,[16] these findings may provide further evidence of the beneficial effects of PA on mental health and well-being.

It is commonly reported that females engage in less moderate- and vigorous-intensity PA than their male counterparts.[17] In accordance, the findings of our study show that in the pre-COVID-19 period, a lower proportion of females (73%) met recommended PA guidelines compared to males (81%). During COVID-19 restrictions, females engaged in less high-intensity PA (e.g., running, cycling, resistance exercises) than males, but more low-intensity activities (walking; yoga/Pilates; Supplementary Table 2). More positive changes in exercise behaviour were shown between pre- and during COVID-19 restrictions for females compared to males. Specifically, in females, the largest increases were found for online exercise classes (0.4% vs. 21.2%, respectively) and online yoga/Pilates classes (0.1% vs. 8.2%, respectively). In contrast, for males, online exercise classes increased from 0.1% to 6.5% and from 0.0% to 1.5% for online yoga/Pilates classes (for pre- and during COVID-19 restrictions, respectively). Self-efficacy, social support, and motivation are empirically substantiated factors that impact on PA levels among women more than men.[18] Little is known, however, about the impact of this pandemic on these factors or even whether such influential factors are altered during a pandemic. As PA is of paramount importance to public health, our longitudinal design will provide data to help explore the barriers, facilitators and adherence to PA, for both females and males, during COVID-19 recovery.

Individuals aged 18-29 years reported the largest negative change (26.1%) in PA behaviour between pre- and during COVID-19 restrictions for all age groups assessed. Previous research has shown that individuals aged 16-34 years typically engage in more aerobic, strength, and sporting activities than people of an older age.[19] In the current study, 18-29 year olds engaged in less resistance-based exercise (35.2 % vs. 19.4% for pre- and during COVID-19 restrictions, respectively) and sporting activities (23.8% vs. 3.6% for pre- and during COVID-19 restrictions, respectively), most likely due to the closure of gyms/fitness centres and the cancellation of all structured team and individual sporting activities (Figure 2; Supplementary Table 3). The decline in participation aligns with the ‘relapse’ stage of the Transtheoretical Model of Behaviour Change. As it has been shown that re-commencing a previously broken PA habit can be challenging, the changes observed in this study when extrapolated to the general population could be detrimental to long-term public health.

Physical activity is an important preventive and therapeutic intervention for a wide range of chronic diseases/conditions. In our study, 17.8% of individuals with a self-reported chronic condition reported a positive change in their exercise behaviour between pre- and during early COVID-19 restrictions. Increases in PA may help mitigate the effects of COVID-19 on this subgroup of ‘higher risk’ individuals by boosting immune function, which is vital to control and eliminate COVID-19,[20] and counteract prevalent comorbidities such as obesity, diabetes, hypertension and vascular conditions.[2, 3] However, 17.3% of individuals with a self-reported chronic condition reported a negative change in their exercise behaviour. Indeed, a negative change may promote the development and/or progression of many chronic diseases, which may contribute to potentially poorer outcomes in those who contract COVID-19.[2] Accordingly, individuals with comorbidities are an important group to consider when designing and delivering guideline recommendations to encourage PA during periods of physical distancing and self-isolation.

#### Effect of country on PA, mental health and well-being

The World Health Organisation reports a higher prevalence of depressive and anxiety disorders in NZ (5.4%, 7.3%, respectively) and AUS (5.9%, 7.0%, respectively), compared to the UK (4.5%, 4.2%, respectively) or IRE (4.8%, 6.3%, respectively).[21] In this study, however, the NZ population demonstrated better mental health and well-being during COVID-19 restrictions than all other countries surveyed. When comparing the two largest survey populations, depression, anxiety and stress were lower in NZ than the UK by 22%, 41% and 24%, respectively. Although there were no differences in PA during early COVID-19 restrictions between these two countries, or indeed any of the countries surveyed, there were differences in exercise behaviour change between pre- and during COVID-19 restrictions. A greater proportion of the NZ study population maintained their pre-COVID-19 exercise behaviour (72.2%) compared to the UK, IRE or AUS (63.7%, 65.3% and 64.2%, respectively). Furthermore, NZ demonstrated statistically fewer negative changes in exercise behaviour (12.6%) compared to the other countries surveyed (UK: 21.3%; IRE: 17.7%; AUS: 22.6%). It is widely accepted that PA is associated with a reduced risk of depression and anxiety.[22] A recent study in the USA demonstrated reduced PA and increased screen time during the early COVID-19 restriction period were frequently associated with poorer mental health outcomes.[4] Similarly, our correlational findings demonstrated that increased sitting times were associated with poorer mental health and well-being. IRE reported the lowest daily sitting time but comparable PA levels to other countries, suggesting that participants in IRE may have been undertaking greater incidental PA. Despite incidental PA being suggested to have numerous practical and physiological health benefits,[23] as well as potential to improve mood and well-being,[24] in this study, IRE reported statistically poorer mental health compared to the other countries surveyed. It is plausible that the COVID-19 pandemic adds additional complexity to such a relationship, and further research into incidental PA and mental health is warranted.

#### Implications of study findings

In countries where physical distancing (e.g., working from home) is likely to feature to a greater or lesser extent in the short-to medium-term, the potential impact of PA and changes in exercise behaviour on mental health and well-being is significant. Marginalizing PA during these uncertain times could have paramount negative implications for public health and thus attention must be paid to helping to promote and support people to engage in PA. Differing health promotion strategies may be required to facilitate engagement from specific groups (e.g., males, younger adults and those with comorbidities). These findings have important implications for policy and guideline recommendations and may assist in refining government strategies concerning physical distancing and self-isolation.

## Conclusion

During early COVID-19 restrictions, a negative change in exercise behaviour compared to pre-COVID-19 restrictions was associated with poorer mental health and wellbeing. Whilst females reported more positive changes in exercise behaviour, young people (18-29 y) reported more negative changes. PA was comparable between the UK, NZ, IRE and AUS, however, people in NZ reported better mental health and well-being. Our findings will assist in the development of targeted interventions to encourage greater PA participation while individuals continue to physical distance, self-isolate, or ‘work from home’ for extended periods. Accordingly, this may positively impact upon individuals’ mental health and well-being during this challenging time. Due to the uncertainty surrounding the long-term effects of the COVID-19 pandemic, longitudinal studies are needed to explore the relationships between PA and mental health and well-being.

## Data Availability

Authors confirm that they are willing to share data upon request.

## Author Contributors

JF, DL, JB, W’OB, BM and DW designed the study. All authors contributed to survey dissemination and preliminary data preparation. BS analysed the data. JF and DL wrote the main draft of the manuscript and all authors contributed to manuscript revision and approved the final version.

## Transparency statement

JF confirms that the manuscript is an honest, accurate, and transparent account of the study being reported; that no important aspects of the study have been omitted; and that any discrepancies from the study as originally planned (and, if relevant, registered) have been explained.

## Data Sharing

Data are available upon request.

